# Can Place-Based Modifications Make a Difference to Local Health Inequalities in Urban Essex: An Evaluation Protocol

**DOI:** 10.1101/2024.08.29.24312816

**Authors:** K. Cusimano, P. Freeman, A. Pettican, A. J. Brinkley

**Affiliations:** School of Sport, Rehabilitation and Exercise Sciences, University of Essex, CO4 3SQ; School of Health and Social Care, University of Essex, CO4 3SQ

**Keywords:** Effectiveness, multiple-deprivation, interventions, implementation, systems

## Abstract

Stemming from a complex picture of compositional, contextual and wider determinants, health inequalities are presented at the level in which people reside (i.e., their place). Examples of this exist within Essex, England, where despite seeming affluence, pockets of high multiple deprivation exist. Programmes delivered across the system representing each place may provide a solution to these complex challenges. For this reason, Epping Forest District Council commissioned the evaluation of a programme representing two place-based projects within their district (i.e., Limes Farm and Oakwood Hill). This paper provides the evaluation protocol for this programme. Broadly, the evaluation seeks to investigate the design, implementation, mechanisms and effectiveness of both projects. Our evaluation is underpinned by the Medical Research Council (MRC) guidelines for the design, evaluation and implementation for complex interventions, and takes inspiration from a realist approach. We aim to understand *where* each project works, *who* does the projects work for, *what* impact do the projects have, and *how* and *why* does the projects work. This will be achieved through a mixed-methods approach which utilises a cohort study, ripple-effects mapping, focus groups, and secondary data analysis. Quantitative data will be analysed using descriptive, general linear and multi- level models, while qualitative data will be understood using visualisation (ripple-effects maps) and reflexive thematic analysis. Data will be triangulated to create programme theory configurations, which explain the outcomes which stemming from the programme, and how these are shaped by mechanisms within a given context. We anticipate our novel and robust approach to contribute to policy surrounding the adoption and implementation of place-based approaches.

## Background

Health inequalities are typically manifested within the *‘place’* in which people reside^1^. For example, life expectancy differs at a country (i.e., England; 78.8 years - men, 82.8 years - women), county (i.e., Essex; 80.3 years - men, 84.1 years – women), local authority (i.e., Epping Forest District; 80.1 years - men, 83.5 years – women), and ward-level (i.e., Grange Hill, Epping Forest; 79.6 years - men, 82.7 years – women)^2^. With significant variation existing between wards of the same local-authority (i.e., 11.8 year – men, 12.7 year - women) in Epping Forest^2^, such as Loughton Roding (i.e., 83.4 years – men, 83.5 years women) and Loughton Forest (i.e., 85.2 years – men, 87.8 years women).

These *‘place-based’* inequalities, along with additional health conditions and complications, are shaped by a complex interplay between compositional (individual-level), contextual (environmental-level), and wider (systems-level) determinants of health^1^. These include but are not limited to: individual predictors of behaviour (e.g., fear, anxiety, competence, experience), multiple forms of deprivation, access and opportunity for services, transport, childcare, housing and education, crime, environmental factors, and policy, rules and regulation^1^. In Epping Forest, Essex, England, this complex relationship between health inequalities, the place, and determinants is no different.

Indeed, across specific wards of Epping Forest determinants of health such as multiple deprivation, low household income, local criminality, opportunity and uptake of education, skills, training and employment, service disparities, housing, and environment exist to differing degrees. More specifically, Limes Farm, Chigwell, and Oakwood Hill, Loughton in Essex, England are two examples of this^3^. Both are nested amongst some of the least deprived wards (10-30%) in England, yet Limes Farm is ranked in the 30% most deprived (20% in income deprivation), and Oakwood Hill ranked in the 20% most deprived (10% in education, skills and training) wards in England^3^. These factors drive health inequalities within individual wards^1^.

More specifically, data drawn from regional public health intelligence data^4^ indicates compared to the Epping Forest District Local Authority, areas within Limes Farm and Oakwood Hill have significantly more (+.2-4.2%) individuals living with long-term disability or illness when compared to the local authority average (15.7%). Moreover, these data also indicate a +25.6 difference in standardised incidence ratio for all cancers, and +19.8 difference in standardised admission ratio for emergency hospital visits^4^ for the wards facing the greatest inequalities when compared to the local authority average.

There is evidence which suggests determinants of health inequalities have the potential to be addressed by system-based approaches applied to a given place^1^ ^5–10^. Tailored to the specific needs of each place, and represented by a range of philosophies (e.g., whole-systems), these approaches broadly bring together a range of the right partners, stakeholders, and organisations to identify, explore, and come together^11^ around collective missions and interventions in a place (e.g., improving cardiorespiratory health, reducing alcohol consumption, smoking cessation, employability)^1^ ^5–10^. An umbrella review of systematic reviews on the effectiveness of high- and low-agency place-based approaches across public health and health inequalities broadly found that programmes modifying a range of compositional, contextual, and wider determinants of health such as housing, public spaces and the environment, amenities, active transport, and physical activity opportunities^1^. However, these studies varied in methodological quality^1^ ^5^. In particular^12^, there remains a paucity of evidence encompassing the implementation of place-based programmes, their long-term effectiveness, and intended and unintended outcomes^5^.

Through 2022 and 2023, a place-based project was trialled within the multiple-deprived locality of Ninefields, Epping Forest (see attachment one for the evaluation report). This project adopted a whole-systems^13^ ^14^ approach to address potential determinants of place-based health inequalities (i.e., better access to mental health support, transport, financial and employment advice, digital inclusion, and improved community safety, needs and service awareness), and was supported by in the region of £500,000 of funding. Across the integrated care system (i.e., a partnership approach combining local and regional health and care organisations, upper-tier local councils, the voluntary sector, social care providers, universities, charities, and other partners to collaborate, commission and implement joint health and wellbeing services), upwards of 20-partners engaged >1000 residents in projects and interventions. This included: increased local staffing, provision, and resources; community safety events; tailored physical activity; mental health and weight management programmes; youth clubs; cookery and life skills workshops; financial and employability advice; a bereavement café; clinical investments; and changed natural and built environments. Whilst some interventions were intended to modify the environments individuals reside within (e.g., community safety), many of the interventions were based within a community hub. Recently, this approach has been scaled-out across Epping Forest into Limes Farm, Chigwell, and Oakwood Hill, Loughton. With an evaluation of each project location’s complexity, effectiveness, and implementation being commissioned.

This is important, because, notwithstanding the reach of the pilot, the previous evaluation of this place-based approach had limitations. Indeed, there was the absence of a robust evaluation framework^11^, a means to track long-term within-participant change and the experiences of participants, the extent a systems approach is being implemented, and a method to monitor local- and systems-level intended and unintended inputs, outputs, and impacts of the project^13^ ^15 16^. Addressing these challenges could provide richer and stronger evidence for the effectiveness, feasibility, and implementation of place-based approaches^11^. If such projects prove to be successful, and these are evidenced robustly, this in turn may hold the capacity to lever upstream policy changes across integrated care systems and boards responsible for public health commissioning^17^ ^18^.

The aim of the present study is to understand the design, implementation, mechanisms, and effectiveness of providing place-based projects in two places of high multiple deprivation in Epping Forest District, Essex (i.e., Limes Farm and Oakwood Hill). An overview of our methodology, including our evaluation design and methods is presented below.

## Methodology

### Evaluation Design

The evaluation is underpinned by the MRC guidelines for the design, evaluation, and implementation of complex interventions^15^. These broadly suggest the importance of considering context, programme theory, stakeholders, uncertainty and refinement, and economic considerations across the feasibility, implementation, and evaluation^15^ of complex systems-based interventions^12^. Moreover, this framework encourages the application of mixed-methods^15^ to pick apart this complexity and the intended and unintended elements of complex interventions. To complement this framework, we will take inspiration from a realist philosophy^19^. More specifically, utilising realist questions^20^, the evaluation team will investigate: *where* the project works, *who* does the project work for, *what* impact does the project have, and *how* and *why* does the project work. The answers to these simple questions provide robust data central to the design, implementation, and evaluation of complex systems-based interventions^15^ ^19^. Alongside support from the project steering group, our research team will use these data to generate a programme theory (CMO)^19^. For each place, and as a combined project, this will outline how outcomes (O) (e.g., changes in mental health) are modified by intervention mechanisms (M) (e.g., community gardening) under given contextual (C) settings (e.g., deprivation). The project has received institutional ethical approval (ERAMS: ETH2324-1384) and has been registered on the Open Science Framework (osf.io/pnyrg).

### Overview of the ‘Health and Wellbeing’ Programme, its Projects and Interventions

The delivery of each place-based project is based upon a programme logic model (see Figure 1) developed by Epping Forest District Council in 2023. This logic model was grounded on previous place-based approaches^1^, the wider determinants of health model^21^ ^22^, insight drawn from the Ninefields pilot project, and input from systems-partners. The logic model was designed with the aim of identifying vulnerability and understanding the contextual wider determinants of health within each place and reacting to these challenges through a series of place-based interventions.

**Figure 1:**
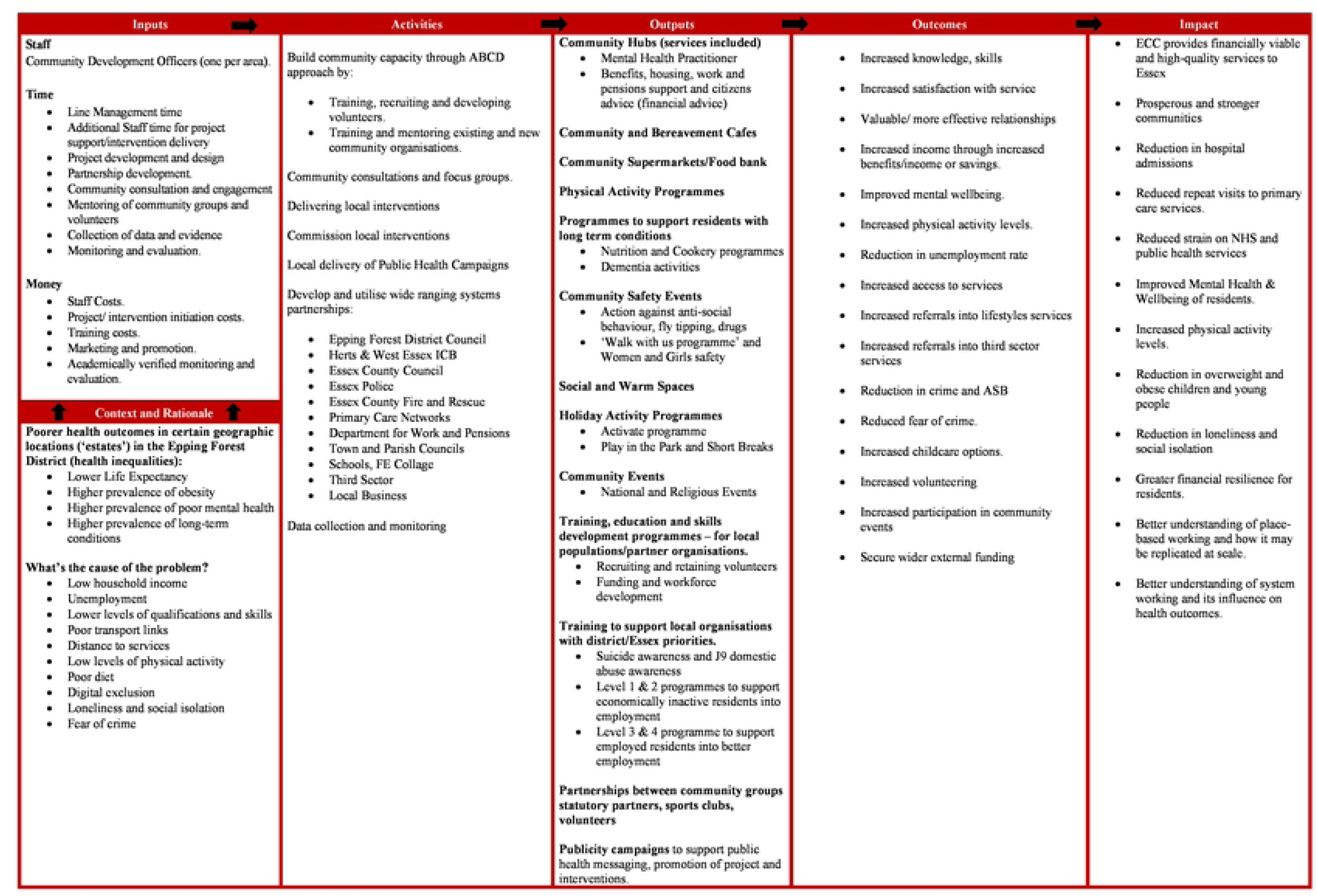
**Epping Forest District Place-Based Health and Wellbeing Interventions Logic Model**

These interventions are the product of partnerships formed across the system, and broadly follow a systems-approach to addressing public health complications^11^. Interventions (outputs) or mechanisms designed to contribute towards outcomes and impact in each place-based context^19^ ^20^ include community hubs delivering tailored support services, community cafes, warm spaces and supermarkets, programmes designed to address health behaviours (e.g., physical activity and dietary behaviours), community safety events, holiday schemes, training, education and employability initiatives, and awareness campaigns. Systems-based programmes in a place are messy, non-linear and complex, and therefore we anticipate a range of unexpected consequences and outcomes stemming from each project^1^ ^5–10^. These will be captured via our methodology and visualised through our programme theory^19^ ^20^.

### Population, Participants, and Recruitment

The programme is focused on two places of high multiple-deprivation in the Epping Forest District of Essex, England^3^ (i.e., Projects in Limes Farm, Chigwell; and Oakwood Hill, Loughton). Within the evaluation, we will sample residents living in these localities, and programme partners, stakeholders, deliverers, and community representatives seeking to effect change in these areas.

#### Population and Participants – Residents (Limes Farm and Oakwood Hill)

The population of Limes Farm is 3,800 people. The area is ethnically diverse (i.e., 61% White British), with 25% of residents aged <19-39 years and 25% of residents aged 39- to 45 years. Across Limes Farm, 37% of households are deprived in one dimension, 14% in two dimensions, and 4% in three dimensions^3^. Health and disability vary within the locality, with only 48% reporting good health (compared to 51.8% within the local authority) and 17% reporting a disability^4^. Moreover, more than one-third of the adult population are economically inactive (37%), whereby 55% have not worked in 12-months, and 31% have never worked.

One thousand one hundred residents live in Oakwood Hill. The population is younger, with 32% of residents aged <19 years, and 26% aged 25- to 39 years, and less ethnically diverse (85% White British). Within Oakwood Hill, 35% of households are deprived in one dimension, 24% in two dimensions, and 7% in three^3^. Health status is similar to Limes Farm (47% report good health), however, 5% report very bad health (compared to 1% within the local authority)^4^. Within the locality, 18% have a disability (compared to 5.8% in the local authority). Economic activity is low, with 39% being economically inactive, of which 62% have not worked in the last 12-months and 27% have never worked.

Based upon the pilot conducted within Ninefields, as a total we anticipate upwards of 3,000 residents engaging in the place-based project (i.e., 2,500 in Limes Farm, and 500 in Oakwood Hill). An overview of our inclusion and exclusion criteria for participants in the evaluation are outlined below. Our inclusion and exclusion criteria are unassuming whereby participants will be included if they are over 18 years of age, and reside in either Limes Farm, Chigwell or Oakwood Hill, Loughton at the time of sampling. Participants will be excluded from the study if they unable to provide informed consent.

#### Population and Participants – Partners, Stakeholders, Deliverers and Representatives

The projects in each place are supported by programme partners, stakeholders, deliverers, and community representatives across the integrated care system. Central to these is Epping Forest District Council who coordinate and fund the project. The council are supported by range of partners, such as the Hertfordshire and West Integrated Care System and Board, Essex County Council, Essex Police, Essex County Fire and Rescue, the primary care networks in the region, the Department for Work and Pensions, the town and parish councils which represent the localities, third sector organisations, schools, further education colleges and universities, and local business. Participants from these organisations will be included if they are over the age of 18 years, and excluded if they are unable to provide informed consent. We anticipate the number of organisations involved within the project will evolve as a function of: (i) changes in place-based needs, (ii) funding, (iii) awareness of the project, and (iv) integrated care system policy.

#### Recruitment

Sample size and sampling strategies are outlined within each methodology. Broadly we will use a multi-method approach to recruit residents into our cohort study and/or our focus groups. Indeed, in the cases of our cohort study, when visiting a project hub or intervention for the first-time participants will be encouraged to complete the survey. To capture individuals who are impacted by place-based modifications, such as infrastructure change, but do not enter a hub or are involved in an intervention directly, we will utilise posters, business cards, and advertisement (to be agreed) with QR codes to a sign-up website across each locality. In the case of our focus groups, we will use these approaches, alongside purposeful recruitment to capture a heterogeneity of individuals residing across each place. Within our ripple-effects mapping (REM) workshops with programme partners, stakeholders, deliverers, and community representatives’ participants will be recruited through email invite.

### Data Collection Methods

#### Cohort Study

##### Design

The evaluation of the effectiveness of placed-based projects presents meaningful challenges to traditional designs such as the randomised control trial^23^. In our case, these include resistance to randomisation by participant or stakeholders (e.g., acceptability), contamination (e.g., participants engage with multiple interventions within and external to the place), feasibility of randomisation, or non-adherence to control group allocation^23^ ^24^. In addressing these challenges, we will examine the effectiveness of each place-based project using a longitudinal cohort study. Observational studies where a cohort of participants are monitored over time are widely accepted to be an alternative where randomisation is deemed not acceptable or feasible within place-based evaluations^25–28^. Within our study, we will assess a range of health, wellbeing, and societal outcomes through an online survey (See Attachment 2 for a complete overview) delivered via Qualtrics™ delivered at baseline (T^0^), and then at 3- (T^1^), 6- (T^2^),12- (T^3^) and 18-month (T^4^) follow-up intervals. Residents will be invited to participate in the study through their first interaction with any project intervention (e.g., attendance at a community safety event).

##### Participants and Sampling

The nature of place-based projects is that they are reactionary to needs of the place^1 11^. For this reason, when calculating sample size, utilising exact existing effect sizes drawn from previous literature prove little use, as the nature of the interventions implemented (i) vary significantly and (ii) are context and systems driven. Therefore, we adopted a recent umbrella review of systematic reviews of existing place-based interventions^1^ to estimate the potential effect of the modification observed in each place. This indicates changes across the built and physical environment, infrastructure, housing, homes, public realm (e.g., environmental changes), supermarkets, transportation, and economy, and via multi-component change small to medium are likely to be observed within our outcomes^1^. We therefore anticipate observing small (*d*=.20) to medium (*d*=.50) effects across the study outcomes.

We plan to utilise longitudinal growth curve modelling^29^. Based upon artificial data modelling^30^, an estimated small effect (*d*=.20), five time-points, *^1^-β* =.95, and *α*=.05, 550 participants (2,750 units of observation) will be required to observe a small effect in our outcome measures. Multi-level models are sufficiently robust to deal with missing data^29^, offer superior statistical power to general linear models, and robust to control for confounding variables (e.g., time). However, given anticipated high levels of attrition, we will inflate our sample size by 30% (715 participants; 3575 units of observation).

#### Measures

##### Demographic Markers

To assess inter-individual variation in intra-individual responses age, gender, ethnicity, disability/long-term health conditions, dependency, postcode (as a proxy measure for multiple-deprivation)^3^ will be requested from participants.

##### Intervention Dose

To understand dose across each place-based project and their interventions (e.g., engagement with a physical activity intervention) participants will be asked a self-report questions on engagement (i.e., *how long have you been using the health and wellbeing hub or services they provide* – Never to 2 years or more) and frequency of attendance (i.e., *on average how often do you attend the health and wellbeing hub or use the services they provide –* less than once a month to daily). Given each project responds to place-based health inequalities, we will adapt these questions as the project evolves to include multi-response items for intervention options.

##### Physical Activity Behaviour

The International Physical Activity Questionnaire Short Form (IPAQ)^31^ will be used to provide a self-reported estimate of physical activity behaviour. The IPAQ is a widely utilised measure of self-reported physical activity behaviour^32^ and examines the frequency and dose of vigorous and moderate activity, walking, and sitting, using seven items^31^. Using the standardised scoring protocol^31^, items are used to calculate metabolic equivalent of task per-week, as total for each domain of activity, and as a total sum. Whilst a systematic review of the IPAQ’s application has reported a poor correlation with objective measures^32^, indicating over- and under-estimation, the measure does present an acceptable, feasible and non-invasive methods to assess physical activity within the present study^31^. Moreover, the measure provides more detail on intensity than alternatives such as the Single Item of Physical Activity^33^, and presents meaningfully less burden on the participant than the recall diary adopted within the Active Lives Survey^34^.

##### Lifestyle and Nutrition Battery

Whilst multiple healthy lifestyle and nutritional questionnaires exist, these range in duration and population specificity. To complement our existing measure of physical activity and sedentary behaviour, and build picture of a healthy lifestyle^35^, we will examine smoking, alcohol and food behaviours, and body-mass index (BMI). In doing this, we designed and will adopt a self-reported battery. This includes categorical response (e.g., yes, have in the past, no) and continuous response questions representing smoking and vaping (i.e., number of cigarettes/e-cigarette/vape), alcohol consumption (i.e., units of alcohol), fruit and vegetable consumption (none to >6 servings), and dietary sugar content (none to >6 servings)). To assess BMI (kg/m^2^) we will request participants provide self-reported height (cm) and weight (kg).

##### Quality of Life

To understand quality of life, we will adopt the Dartmouth COOP Functional Assessment Chart (COOP)^36^. The COOP provides an indicator of quality of life across nine dimensions (i.e., feelings, social activity, changes in health status, physical fitness, pain, social support, health, daily activity, and general quality of life) across a 4-week period on a 5-point Likert scale (1= not at all, 5 = extremely). The COOP has demonstrated good acceptability and feasibility in a UK community population^37^, good psychometric properties^36^, and acceptable intra-class correlation coefficients across all items (.17-61) over time^38^

##### Loneliness

The UCLA loneliness scale (Version 3)^39^ will be used to chart loneliness. Across eight items and on a 4-point Likert Scale (1 = never, 4 = often), participants will rate the extent eight statements reflect their perceptions of loneliness^39^. The measure holds good re-test reliability (*r*=.73) and a strong Cronbach Alpha (*α*=.89-.94)^39^.

##### Mental Health and Wellbeing

Mental health and wellbeing will be understood using the Short Warwick and Edinburgh Mental Wellbeing Scale (SWEMWBS)^40^. The SWEMWBS invites participants to respond to 7-items on their perceptions of their mental health and wellbeing^40^ on a 5-point Likert scale (1 = none of the time, 5 = all of the time). Scores are transformed following measure guidelines (see Stewart-Brown et al^41^ for an overview), whereby higher scores indicate greater mental health and wellbeing. The measure has demonstrated excellent psychometric properties in a range of clinical and public-health settings^40^, and holds the capacity to demonstrate a responsiveness to change^42^.

##### Community Trust, Financial Stability, Disposable Income and Health Service Access

In addition, several items will be adopted from the national-level longitudinal studies. These include an item for community trust from the Active Lives Survey^34^ (‘to what extent do you agree or disagree that most people in your local area can be trusted?’), where participants respond on a 5-point Likert scale (1 = strong disagree, 5 = strongly agree). Moreover, we will include items from the Understanding Society Survey^43^, such as two questions relating to financial stability (How well would you say you yourself are managing financially these days? – responded to on a 5-point Likert scale from 1 = finding it quite difficult to 5 = living comfortably) and reported disposable household income, and an item whereby participants report visits to their GP or family doctor in the past 12- months.

### Ripple-Effects Mapping

Complex systems place-based projects typically hold intended and unintended consequences for participants, stakeholders, and partnerships, and are shrouded by a myriad of enablers and challenges to implementation^5^ ^12 44^. Whilst longitudinal studies can provide good evidence on effectiveness, they are limited to the measures observed. Typically, these reflect what the intended outcomes derived from participating in each project or intervention are^44^ ^45^. However, complex projects, and particularly those modifying wider systems-level determinants are modified and result in unintended impact, consequences, and complications^5^ ^12 44 45^. For example, an active transport scheme creating new cycle paths may intend to improve physical activity behaviour, but unintendingly benefit the local economy (e.g., use of local bike shops), lead to the creation of inclusive local cycling clubs, reduce CO_2_ emissions, and reduce the incidence of road-traffic collisions. These ripple-effects are common within every complex intervention mechanism and can be understood through REM^12^ ^44 45^.

Through workshops, REM understands if changes in an outcome are the product of a project intervention (e.g., community safety), a series of broader contextual factors, the system’s dynamics, and broader unintended social, behavioural, health, and wellbeing values of participation^44^ ^45^. Typically, this is conducted along a timeline, where participants retrospectively trace the history of a project, and using appreciative inquiry (i.e., thinking positively) envisage its future^44^. Moreover, along this timeline participants document the benefits stemming from each project, the extent these were intended or unintended, and the extent to which these were impacted by contextual and systems factors and an array of determinants^44^. This methodology has grown in popularity in recent years, with robust examples in the domains of physical activity promotion^44^, education provision^46^, nutrition provision and education^47^, and place-based wellbeing programmes^48^.

Within our approach, programme partners, stakeholders, deliverers, and community representatives (*n*=15) from each place will be invited to participate in four independent half-day workshops (i.e., approximately every 6-months) (i.e., *n*=8 workshops). Participants will represent a heterogeneity of roles and responsibilities across the system and place. Along the timeline, participants will be invited to consider how this impact has been shaped by factors internal and external to the broader programme, the extent to which changes were intended or unintended, the most and least significant forms of impact, and if these changes represent a broader trend of changes within their place. This process will be tailored to each participant group to explore how these outcomes are maintained, modified, adapted, or lost throughout the project. This sequential process will be completed with the same group of participants at each timepoint. Building on recent advances in the methodology^49^, we will ask *‘realist’* style questions to examine the how, what, when, where, and why of each ripple-effect. The responses to these questions will be documented using a voice recorder. Using Miro™, analysis will produce a visual timeline outlining each intended and unintended *‘impact’* identified. Data from recording will be transcribed verbatim.

### Focus Groups

Understanding the experiences of users within complex interventions is vital in exploring the contextual and inter-individual variations in experience stemming from implementing place-based modification^50^. Additionally, it may reveal incidences where complex systems place-based interventions have unintentionally compounded and reinforced existing health inequalities. For example, there may be individuals who have struggled to participate in some interventions due to certain place-based limitations, such as the accessibility of transport links and buildings. To do this we will conduct semi-structured focus groups with residents of each project area (*n*=10, 5 per-place) approximately every 6-months. The exact number of focus groups and residents participating in these will be underpinned by data saturation^51^ ^52^. Accessibility of the form of focus group will be paramount, to ensure the inclusion of a diverse group of residents and experiences. Therefore, the form of focus group will be negotiated with potential participants and may, for example, take place in person or via an online meeting platform such as Zoom, which has been found to be particularly accessible^53^.

We will purposefully sample a heterogeneity of demographic factors and backgrounds, and residents who use or have access to the interventions across each place (e.g., health hub). Recorded via a digital voice recorder or via a platform such as Zoom, the focus groups will present broad questions to a group of approximately 6-8 people, to promote discussion and debate. Our discussion questions will be focused on a series of realist questions^19^. This will broadly focus on *where* the project works, *who* does the project work for, *what* impact does the project have, *how* does the project work, and *why* does the project work. Data will be transcribed verbatim.

### Secondary and Programme Data Collection

To examine the broader long-term implementation and impact of the provision of each place-based project, we will analyse several publicly available datasets (e.g., Office for National Statistics Data, Understanding Society, Active Lives), and programme data collected by Epping Forest District Council. We will use these data to examine questions related to but not limited to the number of residents engaged with each intervention or modification, number of interventions delivered, funding allocated, social and economic return on investment^54^, changes in place-based health, wellbeing, behaviour and perceptions (e.g., of community safety), and number of residents accessing health, clinical, lifestyle, and social services.

### Data Analysis

#### Quantitative Data Analysis

##### Cohort Study

Data drawn from our cohort study will be analysed using longitudinal growth curve models^29^ on MLwiN 3.10. These multi-level models examine longitudinal intraindividual and interindividual variability^29^. The models are sufficiently robust to cope with equivocal missing data, unequally spaced time points, non-normally distributed data, dependent error structures, time-based covariance, heterogenous variance, and moderation effects^29^. To address missing data, we will apply the intention to treat principle^55^, where data missing completely-at-random will be managed using the last-observation carried forward or back. Our model will be constructed across two levels, whereby time- points (level 1) (*n*=5) are nested within individual participants (level 2). Within multi-level models, >15 units are required to form a level of analysis^29^, therefore until interventions are established within each place, it remains unknown if a 3-level model whereby participants would be additional nested into interventions, and subsequent analysis is possible. To any extent, the model will be estimated through Iterative Generalised Least Squares and constructed through a sequential process. This includes establishing an interclass correlation coefficient (variance component *‘null’* model), prior the construction of a fixed (model 1) model where period effects (i.e., fixed predictor of time) will be entered into the model on a grand mean centred basis. Interindividual variation will be examined via a random slope (model 2) model whereby study outcomes will vary as a function of explanatory variables (i.e., demographics). Model fit will be calculated through 2*loglikelihood and χ^2^ distribution tests for significance.

##### Secondary Data Analysis

To investigate questions relating to reach of the project (i.e., absolute numbers, proportion of engagement) using JASP™ (0.18.3) we will utilise a range of descriptive and general linear models. More specifically, we will report descriptive statistics for the number of interventions or modifications delivered, the number of residents engaged in these initiatives, and funding allocated across each project. To examine the variation of participation in (delivered) interventions we will conduct multiple regression to examine the determinants (e.g., age, gender, deprivation) which shape participation.

To examine the impact of each project (e.g., long-term effectiveness) across social, economic, and health markers (and social and economic return on investment^54^ ^56^) and the extent health, clinical, lifestyle, and social services are accessed, where data is historically available and collected on equally spaced intervals (e.g., Active Lives data), we will utilise interrupted time series analysis^28^ on JASP (0.18.3) using the *‘Prophet Model’*. Interrupted time-series analysis examines the change in a historical trend of continuously collected data by an interruption (e.g., typically an intervention, modification, or event)^56^. Within a time series, analysis seeks to examine if a difference exists between the counterfactual trend (i.e., historical trend to date) and present trend (i.e., the one potentially caused by the intervention)^56^. It therefore is particularly helpful in understanding the impact of broader interventions that seek to modify the system as a whole^27^ ^56^.

Within the analysis, we will foremost construct a visual overview of the trend using descriptive statistics and scatter plots. This will highlight the expected trend, and any unexpected outliers and complications with seasonality and time-varying confounders. Following this process, we will utilise a Poisson multiple regression model (count data) or linear multiple regression (continuous data), where an outcome will be predicted by time, and a dummy variable representing the period pre- and post-intervention (i.e., beginning of the place-based projects). To account for residual autocorrelation (if required), we will utilise autoregressive integrated moving average. To address seasonality, a replicable period of data will be included pre- and post-implementation. Whilst typical confounders (e.g., age, gender, multiple-deprivation) are controlled through the population representative nature of the datasets utilised^56^ and stability over-time, we will account for event-changes (e.g., change in national policy, pandemic, weather) via dummy variables within the regression model. In all cases, slope change will be calculated to examine the difference between the counterfactual trend and interrupted trend. In the case of analysis representing health and wellbeing outcomes (cohort study and secondary data analysis), these changes will be used to calculate social and economic return in investment^54^ in accordance with the HM Treasury’s Green Book^57^ (e.g., the ‘wellbeing-adjusted life year’ – WELLBY). The WELLBY provides a simple estimate of how change in wellbeing contributes to social and economic outcomes (i.e., 1-point change equals a median £13,000 increase in societal and economic outcomes such as health and social care needs)^57^.

### Qualitative Data Analysis

Transcribed voice recordings from REM and focus groups, and impact pathways drawn from REM will be analysed using reflexive thematic analysis^58–60^ on NVivo™ (latest version). Foremost, following familiarisation, all data will be coded inductively by the project research associate (KC) to create a long-list of codes within the data. These codes will then be categorised based upon their commonality. Here, initial themes will be generated. Through a process of critical appraisal within the broader research team, these themes will be specified by data, and the coherence of the themes will be evaluated. Within this process, sub-themes will be generated and specified as appropriate^58–60^. These themes will explain how, when, where, why, and for whom the projects work. To remain reflective of our position as outsiders within each project and population^61^, whilst conducting the research we will remain open, non-judgemental and the project research associate will retain a reflexive diary. We anticipate this assisting the research team in addressing positional reflexivity^62^ ^63^ (e.g., shaping themes based on a narrative created through immersion in the programme evaluation alone).

### Triangulating the Analysis and Generating Programme Theory

Using a methods approach^64^ ^65^ (i.e., utilising the strength in each form of data) and inspired by a realist approach^19^ ^20^, quantitative and qualitative analysis will be triangulated to create a programme theory (CMO) explaining the how, what, why, when, and where of each project. In our case, we will produce a programme theory for each place-based project and the overall broader programme theory for the programme. Initial programme theories will be generated by the research team and revised with support of the programme steering group (i.e., a range of partners involved in the delivery and management of the project).

## Discussion

There is growing evidence highlighting the role of the place in health inequalities^1^. Complex interventions implemented within the system representing a place may provide a solution^1^ ^5–10^. The paper provides a protocol for the evaluation of the design, implementation, mechanisms, and effectiveness of two place-based projects in high areas of multiple deprivation in Essex, England. The evaluation is underpinned by the MRC guidelines^15^, inspired by a realist philosophy^19^, and utilises a mixed-methods approach^15^. Within our evaluation, we seek to investigate *where* each project works, *who* does each project work for, *what* impact does each project have, and *how* and *why* does each project work. The answers stemming from these questions will inform the creation of a programme theory (CMO)^19^.

Our protocol serves to contribute to the growing number of papers^12^ ^66^ surrounding the evaluation of complex interventions delivered across the health and wellbeing systems. Whilst elements of our evaluation are fixed, the project we are evaluating is conducted over the long-term. Therefore, we hope, in publishing our protocol, we can garner critical feedback from the broader evaluation community on additional methods we can apply or adapt. The strengths of approach include our cohort study (i.e., assessing within person effectiveness over the long-term), focus groups (i.e., exploring residents’ experiences of change), ripple-effects mapping (i.e., explaining the intended and unintended benefits, determinants and consequence stemming from a place-based project), and secondary data analysis (i.e., explaining impact and social and economic return in investment). Rather than highlighting the deficiencies in previous research, we prefer to add that our work will contribute to a growing body of research within this space^1^.

Moreover, it should be noted, that both our cohort study and ripple-effects mapping process are designed to be sustainable beyond the funded phase of the project. This is particularly important given the impact of place-based projects such as those conducted within Limes Farm and Oakwood Hill are likely to be felt meaningfully in the long-term^1^. Through knowledge exchange activities not limited to scientific presentations and publications, briefs, and working with partners across the integrated care system and wide public health space we hope to create an impact on policy concerning the application of place-based approaches (e.g., The UK Government’s Levelling Up Agenda). Our approach presents a novel and leading approach to evaluate the policy landscape implemented across these places, through providing robust evidence of our approach we hope to have an impact within clinical and public health systems such as regional integrated care systems aiming to address broader public health complications.

## Data Availability

All relevant data from this study will be made available upon study completion.

## Acknowledgements

The present study is a protocol of an evaluation. Therefore, there is no data to present. The research is funded by Epping Forest District Council, as part of the broader Essex Public Health Accelerator Bids Grants Programme Fund (University of Essex RCP: 17188). As a function of a formative approach to evaluation, the funders do play a role in the evaluation design. Indeed, the research team sit within the programme steering group. Rather than biasing the methods, this group provides useful input into the application of the methods into practice.

